# UVESCREEN1: A randomised feasibility study of imaging-based uveitis screening for children with juvenile idiopathic arthritis- Study Protocol

**DOI:** 10.1101/2024.12.12.24318929

**Authors:** Sonali Dave, Jugnoo Rahi, Harry Petrushkin, Ilaria Testi, Dipesh E Patel, Ameenat Lola Solebo

**Author notes:** **Corresponding author:** Ameenat Lola Solebo 0000-0002-8933-586, Postal address: UCL GOS Institute of Child Health, 30 Guilford Street, London, WC1N 1EH, UK.

## Abstract

Children with Juvenile Idiopathic Arthritis (JIA) are at a currently unpredictable risk of a blinding, often asymptomatic, co-existent eye disorder, anterior uveitis which requires prompt treatment. The unpredictability of this insidious disorder commits children with JIA to three-monthly expert clinical examination in specialist eye centres often located far from their homes. Optical coherence tomography of the ocular anterior segment (AS-OCT) has been shown to be an acceptable, repeatable and sensitive modality for uveitis detection, but is not standard care. This feasibility randomised controlled trial (RCT) aims to inform a future full-scale RCT comparing current routine practice (expert slit lamp examination, SLE) to AS-OCT for the surveillance of uveitis in children at risk.

Eighty children aged between 2 and 12 years old and diagnosed with JIA within the preceding year will be included. Participants with an existing diagnosis of uveitis, other ocular co-morbidities or those unable to complete examinations or provide informed assent will be excluded. Participants will be randomised to SLE (control) or AS-OCT (intervention) examination at a frequency consistent with the current national programme for childhood uveitis surveillance. Children in the intervention arm will also have standard examination at 6 and 12 months after study entrance. Outcomes of interest will be feasibility (recruitment and attrition rates), clinical metrics (proportion diagnosed with uveitis or other ocular disorders at 12 months after study entrance), quality of life outcomes (PedsQL), and resource use. Additionally, comparative analysis of AS-OCT versus SLE (‘gold standard’ reference testing) findings at 6 and 12 months for those in the intervention arm will provide the proof-of-concept data necessary to develop and undertake a larger scale trial.

Trial registration: This trial has been registered with clinicaltrails.gov (NCT05984758).

**Summary:** Anterior uveitis is a rare form of eye disease that is commonly associated with Juvenile Idiopathic Arthritis (JIA). Childhood uveitis is potentially blinding, so children with JIA must travel to a specialist centre every two to three months for an eye examination to pick up signs of uveitis. This feasibility study will compare routine Slit Lamp Examination (SLE) with imaging-based surveillance using Optical Coherence Tomography of the Anterior Segment (AS-OCT) to support the design of a future study. Eighty children between the ages of two and twelve years old who have been diagnosed with JIA in the past year will be invited to take part. Children will be randomised into one of two arms (SLE or AS-OCT) and will have their routine uveitis screening appointments over one year using either of the screening modalities. This study will provide the proof-of-concept data necessary to develop and undertake a larger scale trial.

## Introduction

Childhood uveitis is a rare but significant chronic eye condition [1]. Whist childhood onset disease accounts for approximately 1% of cases seen in adult uveitis clinics [2, 3], it accounts for 10% of cases of uveitis related sight impairment amongst adults [4, 5]. This over-representation is compounded by the frequent presence of multisystem disorders amongst those with childhood onset uveitis [6]. A key example is Juvenile Idiopathic Arthritis (JIA), the most common childhood inflammatory arthropathy [7]. Uveitis affecting the front section of the eye (anterior uveitis) the most common extra-articular manifestation of JIA [8] Childhood anterior uveitis can be asymptomatic, or occur in children too young to report symptoms, so children with JIA undergo regular eye surveillance examinations every few months [9].

The strongest predictor of poor outcomes in childhood onset uveitis is the presence of established ocular structural complications at presentation i.e. delayed diagnosis [10]. The worsening international workforce shortfall in paediatric ophthalmology is an obstacle to timely diagnosis for this population [11]. Imaging-based disease surveillance, which could be undertaken in settings across primary to tertiary care [12], would address this shortfall in capacity, providing specialist level care (potentially closer to the child’s home) to facilitate timely diagnosis and reduce the burden of uveitis related visual impairment amongst children and the adults they become.

Optical Coherence Tomography (OCT) machines have been widely adopted in secondary care and primary care settings (including high street opticians), providing high resolution images of eye structures [13]. OCT of the anterior segment (AS-OCT) to detect inflammatory cells in the eye, the hallmark of anterior uveitis, has been shown to be repeatable and highly sensitive for childhood disease [14]. Work has also been undertaken to understand the normative findings in healthy children [15]. The implementation of this potentially transformational innovation needs robust evidence on safety and effectiveness.

We aim to begin to provide the necessary high-level evidence, through a feasibility randomised controlled trial of AS-OCT versus routine clinical examination for the detection of uveitis in children at risk. This proof-of-concept study will compare imaging-based surveillance to the current approach of clinical examination (slit lamp examination-based surveillance) in the detection of uveitis in children with juvenile idiopathic arthritis The proposed study will provide the data needed to develop eligibility criteria, recruitment and retention approaches, to determine the key outcome metrics and study sample size necessary to demonstrate non-inferiority, and to inform the selection of the resource use measures needed to determine cost-effectiveness.

## Materials and Methods

The proposed study has been designed in line with the SPIRIT (Standard Protocol Items: Recommendations for Interventional Trials) guidelines [16].

### Study design

This study will be a two-arm, parallel group, single site feasibility randomised controlled trial (RCT) comparing slit lamp examination-based (SLE, routine care) with AS-OCT based detection of active anterior chamber inflammation over 52 weeks in children undergoing surveillance examinations for juvenile idiopathic associated uveitis. Study participants will be children newly diagnosed (within the preceding 12 months) with JIA. The primary outcomes will be study feasibility metrics (i.e. recruitment and attrition rates). Proof of concept analyses will also be undertaken, with comparison of AS-OCT assessment to ‘gold standard’ clinical examination at 6 and 12 months.

### Setting

This will be a single site study, undertaken within a tertiary/quaternary healthcare setting: Great Ormond Street Hospital (GOSH), England, UK. Participants will be identified and recruited on attendance to routine outpatient appointments.

### Participants

Eligibility criteria comprise:

- New diagnosis (within preceding 12 months) of JIA
- Eligible for uveitis surveillance in line with national guidance (table 1) [9]
- Aged between 2 and 12 years

**Table 1.**
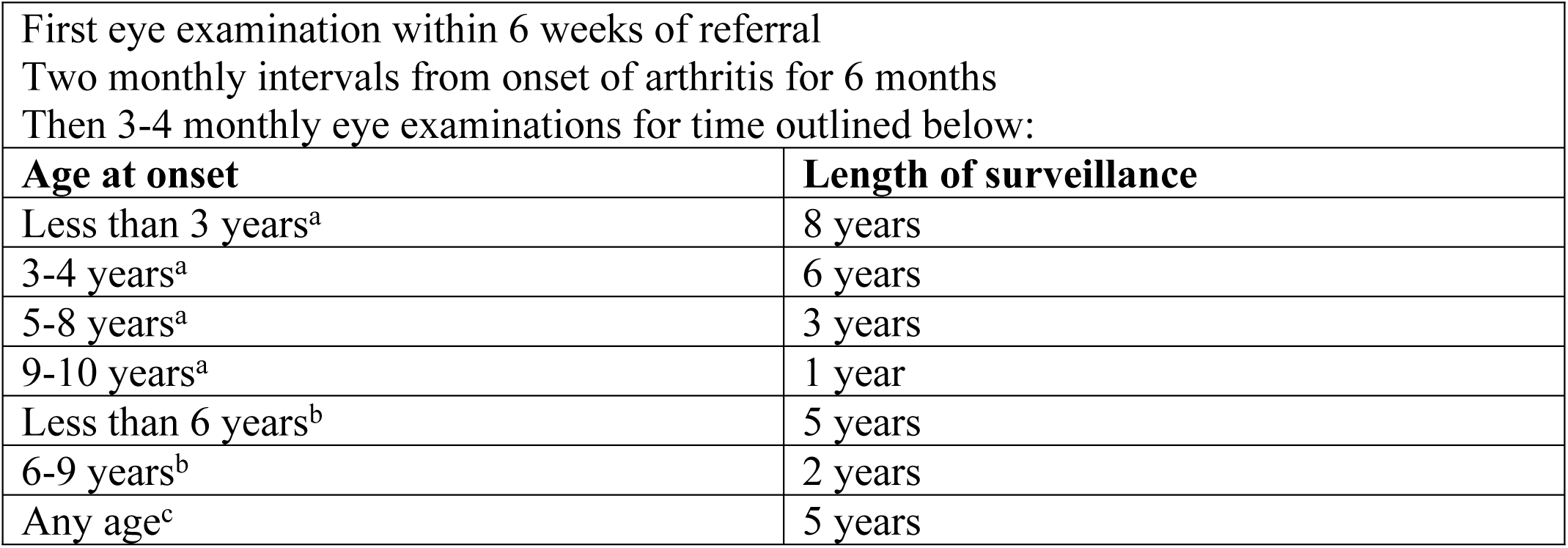
- Guidelines for screening for uveitis in JIA by the Royal College of Ophthalmologists and the British Society for Paediatric and Adolescent Rheumatology (BSPAR). ^a^- For oligoarticular JIA, Psoriatic arthritis onset and Enthesitis related arthritis (ERA) ^b^- For polyarticular, ANA+ JIA onset < 10 years ^c^- For polyarticular, ANA-JIA, onset <7 years

| First eye examination within 6 weeks of referral |  |
| --- | --- |
| Two monthly intervals from onset of arthritis for 6 months |  |
| Then 3-4 monthly eye examinations for time outlined below: |  |
| Age at onset | Length of surveillance |
| Less than 3 years<sup>a</sup> | 8 years |
| 3-4 years<sup>a</sup> | 6 years |
| 5-8 years<sup>a</sup> | 3 years |
| 9-10 years<sup>a</sup> | 1 year |
| Less than 6 years<sup>b</sup> | 5 years |
| 6-9 years<sup>b</sup> | 2 years |
| Any age<sup>c</sup> | 5 years |

Exclusion criteria comprise:

- Visual impairment or any ocular co-morbidities that would negatively impact on best corrected acuity
- Developmental/learning difficulties that preclude concordance with examination/informed assent

### Study intervention - AS-OCT

Children randomised to the intervention arm will undergo AS-OCT imaging to detect anterior chamber inflammation. The AS-OCT images will be acquired using the Optovue RTVue OCT (8 cross-sectional scans) and the Heidelberg Spectralis OCT2 machines (anterior segment cube scan). Images will undergo manual analysis by a trained clinician to identify the presence of inflammatory cells within the anterior chamber. A later semi-automated re-analysis of acquired images will be undertaken within one week of the examination using published protocols [12, 14]. Full details of image acquisition and analysis processes are available in supplemental document 1 (S1).

Imaging will be undertaken at a frequency consistent with line with national guidance (table 1) [9]. This comprises 6 study visits in total (at Month 2 following the first visit to establish eligibility, Month 4, Month 6, Month 9, and Month 12-see table 1 for further details). The full participant schedule of assessments is shown in figure 1, with specific details of the study procedures shown in figure 2.

**Figure 1.**
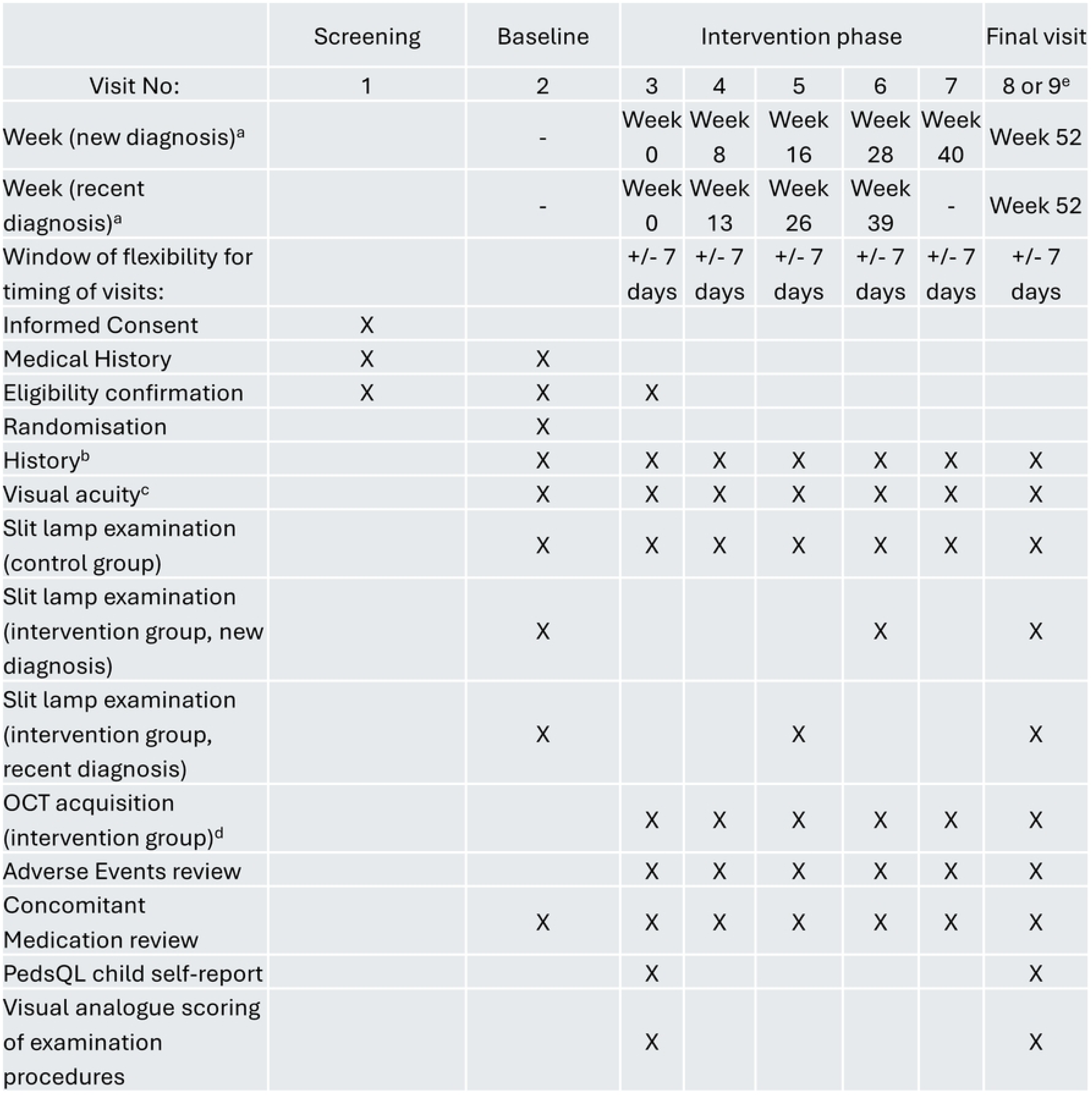
-SPIRIT schedule of assessments. ^a^ Visits for those within 6 months of diagnosis: newly diagnosed: Day 1, Week 8, Week 16, Week 28, Week 40, last visit Week 52 Visits for those entering study outside the 6-month widow: Day 1, Week 13, Week 26, Week 39, Week 52 ^b^ Concomitant use of a. immunosuppression or immunomodulation (and dose); any eye or visual symptoms since last visit; any other parental concerns ^c^ Measured with age-appropriate method: Cardiff cards, Thomson Kays or Thomson LogMAR in each eye separately ^d^ Heidelberg Spectralis OCT2 and Optovue RTVue anterior segment cube and single line scans ^e^ A diagnosis of new onset uveitis at any visit results in exit from the trial

**Figure 2.**
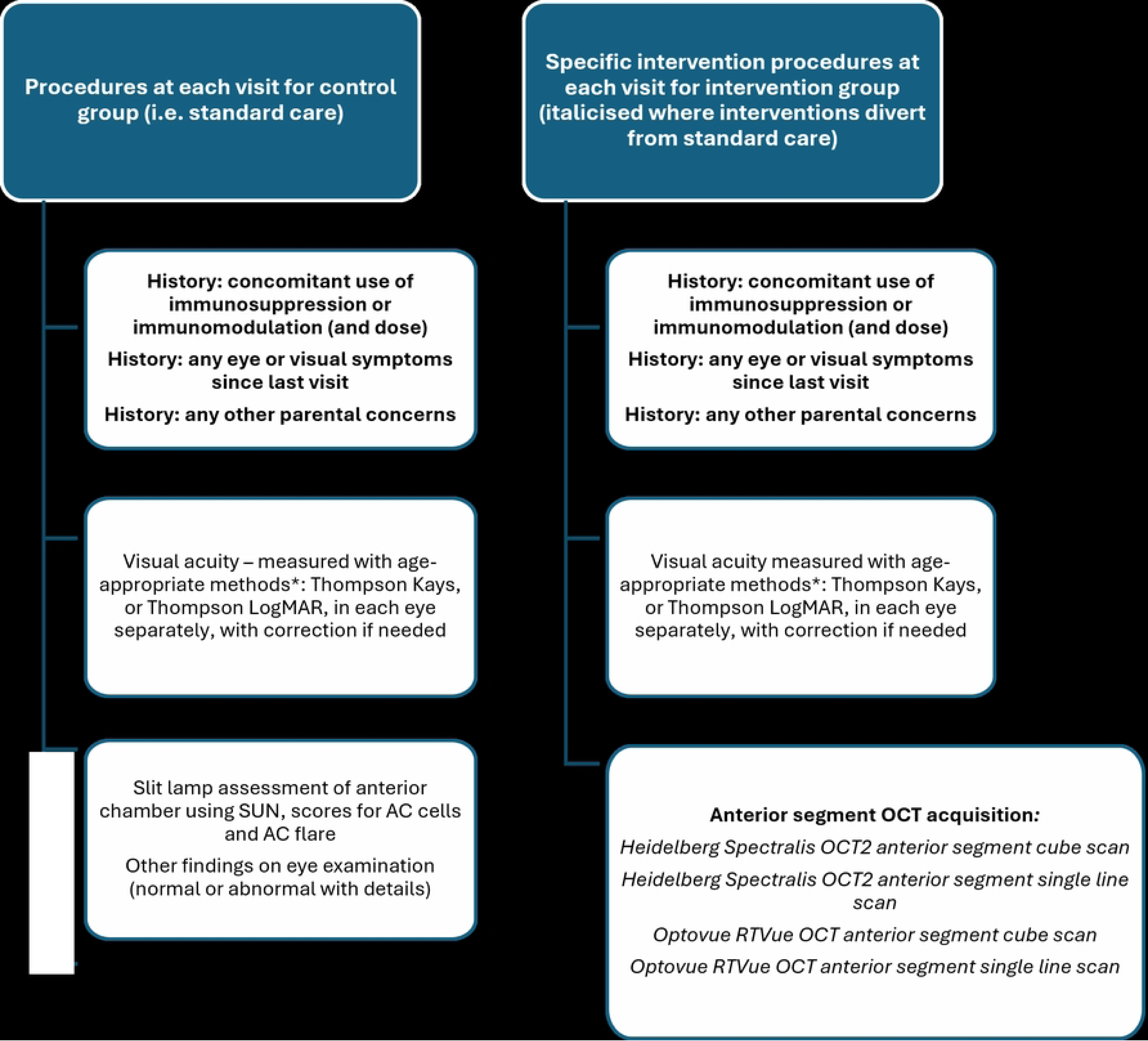
-Details about the specific procedures during the intervention and control arm. *-See supplemental document 2 (S2) for specific visual acuity screening protocol

Exit from the study will occur either at visit 7 (for participants with a new diagnosis of JIA within 6 months of entrance to study), at visit 6 (for participants whose JIA was diagnosed within 6 to 12 months of study entrance, figure 1), or at any visit where uveitis has been diagnosed. A diagnosis of uveitis will be made clinically, using slit lamp examination. Any child within the intervention arm in whom a ‘disease positive’ AS-OCT is acquired (i.e. more than 1 hyper-reflective particle in any cross-sectional scan) will undergo a confirmatory slit lamp examination. Confirmation of uveitis will lead to a child exiting the study.

### Study outcome measures

The main outcomes of interest are the study feasibility metrics, specifically

- The proportion of patients within the service who meet the eligibility criteria, and reasons for exclusion
- The proportion of eligible patients recruited
- The proportion of patients who withdraw due to loss of consent or are lost to follow-up
- Statistical parameters (point estimates and range / variance) of the clinical and patient centred outcome measures

The primary clinical outcome measured (as a surrogate endpoint) will be:

- Agreement, for participants in the intervention arm, of AS-OCT findings with routine clinical examination, analysed at 6 months and 12 months milestones

Other clinical outcomes of interest will be:

- New diagnosis of uveitis
- New diagnosis of other visual/ocular problems

We shall be evaluating patient centred outcomes, specifically

- Health related quality of life (PedsQL child self-report, at 12 months and change from baseline to 12 months)

We shall also be evaluating resource use across the feasibility study to support the design of cost-effectiveness analyses in the larger scale study.

Adverse and serious adverse events (AE/SAE) will be recorded for both the intervention and routine care arms. These events comprise (1) Development of anterior chamber inflammation; (2) development of non-sight threatening sequalae of ocular inflammation during the study period; (3) progression or disease flare of JIA; (4) development of treatment related adverse events. AE/SAE occurring in the intervention arms will be used as criteria to determine progression to a full RCT. Specifically. the progression criteria will be defined using a ‘traffic-light’ system (green – criteria met, amber – minor changes needed, red – significant changes needed).

Recruitment

- Green – >4 recruits/month
- Yellow – 2-4 recruits/month
- Red – <2 recruits/month

Adverse events (incidence of sight threatening complications in imaging-based surveillance group)

- Green / Yellow – No cases
- Red – >1 case

Adherence to surveillance programme

- Green – >75%
- Yellow – 30-75%
- Red – <30%

### Sample size

As a feasibility study, there is no formal power calculation based on clinical non-inferiority for a diagnostic intervention [17]. To ‘quantify’ and assess the concept that imaging-based surveillance is not inferior to clinical slit-lamp examination in the detection of active anterior uveitis in children at risk, the starting point will be that that SLE findings (assumed to be abnormal in 100% of children with an active anterior uveitis) and AS-OCT findings will concord in at least 95%.

### Recruitment

Following identification through medical record review and confirmation of eligibility at routine clinical review, participants will be approached postally, using posted participant information sheets, or by telephone, 1-2 weeks prior to the next planned outpatient attendance. Informed consent will be sought by a member of the research team within a week of being given the study documentation. Assent will also be sought from participating children where appropriate [18, 19] (see supplemental material 3 (S3) for the consent and assent forms to be used in the study). To minimise participant attrition, after entering the trial families will receive telephone calls a few days ahead of each visit to support attendance, and financial support for travel costs.

### Randomisation and allocation

Coordinated registration and allocation of participant trial numbers will be used. Participants will be allocated using computer-generated permuted block randomisation into one of two arms (1:1, block size= 4) stratified by age (8 and under, over 8). This will be undertaken centrally by the coordinating trial team using Sealed envelope (www.sealedenvelope.com). Allocation will not be concealed, and participants cannot be masked to their allocation to routine (slit lamp examination) or imaging-based surveillance (AS-OCT) examinations.

### Data collection and processing

Data will be entered into electronic data collection forms (DCFs) using the REDCap platform, with collection undertaken during study visits by research associates or clinicians and reviewed for completeness by an investigator. Alongside the data collection forms, optical coherence tomography (OCT) images will be collected from participants.

To limit data missingness, the following approaches will be used:

- Validation rules within the REDCap form (i.e. form submission will not be permitted if data entry points have not been completed)
- Incoming data review monthly by the study team and screening committee to identify missingness, and any patterns of / reasons for missingness within the study dataset
- Exclusion from outcomes analysis of participants lacking a full dataset for more than two consecutive study visits

Patient data will not be transferred to any party not identified in this protocol and will not to be processed and/or transferred other than in accordance with the patients’ consent. Direct access to the data will be granted to authorised representatives from the Sponsor, host institution and the regulatory authorities to permit study-related monitoring, audits and inspections. This trial will collect personal data (e.g. participant names), including special category personal data (e.g. participant medical information) and this will be handled in accordance with all applicable data protection legislation. Data (including special category) will only be collected, used and stored if necessary for the trial. The database and randomisation system will be designed to protect patient information in line with the GDPR. All documents will be stored securely and only accessible by study staff and authorised personnel.

### Analysis

Proportions will be reported as point estimates with 95% confidence intervals. Levels of missing data will be explored with respect to certain baseline characteristics, e.g. age and measures of disease severity to assess biases. All harms or unintended effects in each group will be reported. Effect size (of ‘discrimination’ of disease) and variability in outcomes will be estimated with 95% confidence intervals to inform the sample size calculations for a full-scale trial. Initial trial success will be defined as: recruitment rates of >4 recruits/month, or generation of information on barriers to recruitment; adverse event (incidence of new sight threatening complications in imaging-based surveillance group) rate of 0; adherence to surveillance programme of >75% or understanding of barriers to adherence. We will seek to identify potential cost drivers to inform the collection of the data for cost-effectiveness analyses within a future definitive study.

### Data monitoring

This study will be overseen by a study steering committee (SC). The UVESCREEN1 SC will consist of the Chief Investigator, trial staff, and invited members who represent clinical, clinical researcher, and patient stakeholders. The SC will review interim analysis of recruitment figures, adverse and serious adverse events, trial conduct and any substantial amendments to the protocol. The committee will meet 2-3 times a year.

The Chief Investigator will be responsible for the day-to-day monitoring and management of the study. The Chief Investigator will ensure there is adequate monitoring of activities conducted by the study team. This will include adherence to the protocol, procedures for consenting and adequate data quality. The Chief Investigator will inform the Sponsor if there are concerns that have arisen from monitoring activities, and/or if there are problems with oversight/monitoring procedures. Any changes to the protocol including changes to the eligibility criteria, outcomes or analysis will be reported to the study team, sponsor and REC by the chief investigator.

### Ethics considerations and declarations

This study had received ethics approval from Yorkshire & The Humber - Leeds West Research Ethics Committee (REF 23/YH/0237, accepted protocol version V2, date 18/10/2023). The study will be carried out in accordance with the Declaration of Helsinki and all participants will provide written or electronic informed consent.

### Patient and Public Involvement (PPI)

The development of this study has been supported by a patient expert group, the Childhood Uveitis Study steering group [20, 21]. This proposal is also driven by the priorities identified by stakeholders (patients and professional groups) who participated in the 2013 James Lind Alliance Priority Setting Partnership (JLA PSP) [22].

### Dissemination

We estimate participant recruitment to be completed by April 2026 with data collection to be completed by October 2026. Results will be analysed and disseminated approximately six months later. Findings will be disseminated through journal articles and presentations at both academic and medical conferences. Contributors to (i) the design, conduct, data analysis and interpretation, (ii) writing, (iii) manuscript approval and (iv) accountability for the integrity of the work will, depending on their contribution and journal requirements, will be included by name at the manuscript head. The Uniform Requirements for Manuscripts Submitted to Biomedical Journals (http://www.icmje.org/) will be respected in manuscripts generated by this study. At the end of the trial, after the primary results have been published, the anonymised individual participant data (IPD) and associated documentation (e.g. protocol, statistical analysis plan, annotated blank CRF) will be prepared in order to be shared with external researchers.

## Discussion

This feasibility, proof-of-concept RCT will compare imaging-based surveillance to routine clinical examination in the detection of uveitis in children at risk, providing the information necessary for the design of a future study. This information will comprise feasibility data needed to finalise eligibility criteria, recruitment and retention approaches, and data to determine the key outcome metrics and study sample size necessary to demonstrate non-inferiority, and the resource use measures needed to determine cost-effectiveness. Study findings are intended to inform the development of a full scale RCT to determine whether imaging-based uveitis surveillance is non-inferior to routine surveillance with regards to detection of childhood anterior uveitis in those at risk. Qualitative interviews will also be completed with selected participants (n=10) to establish acceptability amongst participants and families and to aid implementation into practice. Longer term impact will be evaluated through the planned full-scale trial.

Attempts to ensure timely diagnosis of childhood uveitis are frustrated by the worsening shortfall in the number of appropriately trained specialists. In some areas of the United Kingdom, children with juvenile idiopathic arthritis are not accessing ophthalmic care within the recommended time window [23]. Across the UK, there is a workforce crisis in paediatric ophthalmology [11]. This crisis is replicated globally [24]. The wide community adoption of OCT provides a platform for the use of AS-OCT to address some of these challenges. Specifically, the use of AS-OCT detection of active anterior uveitis would release patients from the need to attend hospital eye specialist clinics, as imaging could be undertaken in community eye health centres. The findings from this proposed research project would support the future implementation of hospital and community based anterior segment imaging-based uveitis surveillance for populations at particular risk but would also be the foundation of future imaging-based surveillance at whole population level. They also provide a chance to put children first, with childhood disease driving the science and implementation, and supporting later wider adoption for adult uveitis. Adult disease is common relative to childhood uveitis (incidence 17-52.4 per 100,000 versus incidence 2-20 per 100,000) [25, 26], is a major cause of working age blindness [27] and is the most common cause of attendance to acute eye care services [28].

There are limitations inherent in the design of the proposed study. Firstly, due to the study design, it is not possible for the participants and research team to be masked to the arm to which participants have been allocated, and this may introduce the risk of researcher bias. Equally, due to the nature of the intervention, it is not possible for study participants to be masked to their study arm, which also potentially risks information bias in patient reported outcomes. A pragmatic design, in which each child undertook both standard and interventional examination, with randomisation of which examination would be used for assessment, may be the approach taken for future studies, informed by the findings of this feasibility trial. Secondly, recruitment will only take place at a single site, potentially limiting the generalisability of the data. Additionally, this site is a highly specialised care setting, with participants poorly representative of the ‘average’ patient. Again, as this is a feasibility study, informing a future larger scale study, there will be exploration and description of the representativeness of the recruited patient sample, and reflection of how that compares to what is known about the characterises of the wider population of children at risk. Participants in this study will be reimbursed for their travel and will consent to regular phone calls to support attendance. As this deviates from routine care, it can be argued that this reduces the generalisability of the study. However, the findings from this study will provide a baseline for best achievable attrition rate to as comparison for the full RCT, where generalisability will be considered again.

In conclusion, this study will provide vital information about the feasibility of conducting a larger scale trial on SLE versus AS-OCT uveitis screening examinations for children with JIA. The data collected in this study will also provide valuable insight into the practical considerations needed before implementing this into a larger scale RCT.

## Data Availability

Deidentified research data will be made publicly available when the study is completed and published.

## Authors’ contributions

Conceptualization: ALS

Writing-Original draft and Preparation: ALS and SD

Writing – Review & Editing: ALS, SD, JR, HP, DEP, IT

Methodology: ALS

Visualization: ALS and SD

## Data Availability

No datasets were generated or analysed during the current study.

## Acknowledgements

We thank the Childhood Uveitis (patient expert) Studies steering group for their support.

## Supporting documents

Supplemental material 1: Imaging acquisition protocol

Supplemental material 2: Specific visual acuity screening protocol

Supplemental material 3: Consent and Assent forms

